# Management of Acute Appendicitis in Children during Coronavirus Disease-2019, a Perspective of Pediatric Surgeons from South Asia

**DOI:** 10.1101/2020.10.27.20220442

**Authors:** Md Jafrul Hannan, Mosammat Kohinoor Parveen, Md Mozammel Hoque, Tanvir Kabir Chowdhury, Md Samiul Hasan, Alak Nandy

**Author notes:** Corresponding author: Md. Jafrul Hannan, Apt – B3, House – 72/A, Road – 1, Panchlaish, Chittagong, Bangladesh 4203.

## Abstract

**Background:** Non-operative treatment (NOT) of pediatric appendicitis as opposed to surgery elicits great debate and is potentially influenced by physician preferences. Owing to the effects of the COVID pandemic on healthcare, the practice of NOT has generally increased by necessity and may, in a post-COVID world, change surgeons’ perceptions of NOT.

**Objective:** The objective was to determine whether the use of NOT has increased in usage in South Asia and whether these levels of practice would be sustained after the pandemic subside.

**Methods:** A survey was conducted by pediatric surgeons regarding their position, institute, country, number of appendicitis cases managed, and their mode of treatment between identical time periods in 2019 and 2020 (April 1 to August 31). It also directly posed the question as to whether they would continue with the COVID imposed level of NOT after the effect of pandemic diminishes.

**Results:** A total of 134 responses were collected. A significant increase in the practice of NOT was observed for the entire cohort, although no effect was observed when grouped by country or institute. When grouped by position, seniors increased the practice of NOT the most, while juniors reported the least change. The data suggests that only professors would be inclined to maintain the COVID level of NOT practice after the pandemic.

**Conclusions:** Increased practice of NOT during the COVID pandemic was observed in South Asia, particularly by senior surgeons. Only professors appear inclined to consider maintaining this increased level of practice in the post-COVID world.

## INTRODUCTION

As early as the 1950’s, non-operative management of acute appendicitis in children has been practiced.[1] However, treating pediatric acute appendicitis non-operatively rather than by open or laparoscopic appendectomy still elicits great debate,[2–4] spawned by the results of control trials and observational studies[5–9], reviews[10–13], and meta-analyses.[14–16]The meta-analyses were not in agreement. Kessler et al.[15] determined that the operative treatment group yielded a 98% effectiveness, while the non-operative treatment (NOT) group resulted in a significantly lower 74% effectiveness rate. This contrasts the rather positive findings of Georgios et al.[14] who found a 97% success rate with NOT, and Maita et al.[16], who found a 92% effectiveness with NOT in their meta-analyses. Not surprisingly, the results of individual control trials and studies are similar in their disagreement. Knappen et al.[5] used a prospective cohort study of 45 children aged 7-17 years from multiple institutions to study the success rate of NOT of acute uncomplicated appendicitis (AUA). They found that NOT was effective in 93% of the cases, with only 3 participants requiring readmission for an appendectomy during the follow-up period. Similarly, Svensson et al.[7] also reported very positive findings in their randomized control trial of 50 children between the ages of 5 and 15 years. They demonstrated a 92% success rate with NOT with only a 5% (1 patient) recurrence within the one year follow up period and concluded that NOT was a safe and feasible approach for the treatment of AUA in children. In contrast, these studies are the results of Tanaka et al.[8] and Minneci et al.[6]. Although the former found a 98.7% success rate using NOT, 20.8% of the participants required readmission for AUA within the one year follow up period. The latter found that NOT was effective in 85.7% of the cases at hospitalization in a non-randomized control trial comprising 1068 children. However, this number decreased rather substantially to 62.0% by the one year follow up period. These figures are in fact slightly worse compared to their results from an earlier study[9], where they found NOT to be effective in 89.2% of cases at the 30 days follow-up, decreasing to 75.5% effectiveness after one year. The different approaches certainly need to be discussed with the patient and parents[4], although appendectomy is not without its own set of potential complications.[17] Ultimately, it will likely come down to a discussion with the patients and their families, and probably influenced by the personal opinion of the practicing physician.

However, the year 2020 has provided a unique opportunity in which some of this opinion bias, particularly against NOT, can be partially removed. Coronavirus disease 2019 (COVID-19) caused by severe acute respiratory syndrome coronavirus 2 (SARS-CoV-2) emerged from Wuhan, China in December 2019. As of September 2020, the infection has spread globally, with over 43 million confirmed cases resulting in over 1.1 million deaths worldwide.[18] The response to the pandemic has affected all social and economic aspects of life. However, it has also significantly affected healthcare facilities [19] in several ways, such as reallocation of hospital facilities to accommodate the influx of COVID patients, reducing the number of procedures that would normally be carried out as a result of this re-purposing of rooms and also to maintain social distancing protocols to mitigate the spread of the virus. Nearly all countries (90%) have experienced such disruptions to their health services, with low - and middle-income countries reporting the greatest difficulties.[19] This effect has been particularly strained surgeries in general,[19,20] and thus would have a direct effect on treatment strategies for pediatric AUA. The objective of this study was to determine the effect of the present COVID pandemic on the approaches of care for AUA in Bangladesh, India, Nepal, and Pakistan, and to determine if any permanent changes in opinions of care once the effects of the pandemic have diminished will occur as a result.

## METHODS

The instrument used in this study was a survey (administered via SurveyMonkey) addressed to pediatric surgeons in South Asia, particularly Bangladesh, India, Pakistan, and Nepal, consisting of ten questions pertaining to the position, type of institute, country, number of appendicitis cases managed between April 1 and August 31 in 2019 and 2020, and the number and percentage of those that were treated non-operatively (with responses binned within 10% increments). The final question posed to each pediatric surgeon was whether they would maintain their current COVID imposed level of NOT once the effect of pandemic on the ability to perform surgery subsides. The study was approved by the South Point Hospital, Chittagong, Bangladesh (No.Admn/SPH/190/2020).

The survey response data were analyzed using JMP Software V15.2 (JMP, Cary, South Carolina, USA). Matched pair tests for each respondent were the main tool used and the analysis of these results was performed using a Wilcoxan Signed Rank Test. The analyses were performed ungrouped and grouped by country, type of institute, and position.

## RESULTS

A total of 134 physicians responded to the survey. There were no missing data. Their positions, types of institutes, and country of practice are summarized in Table 1.

**Table 1:**
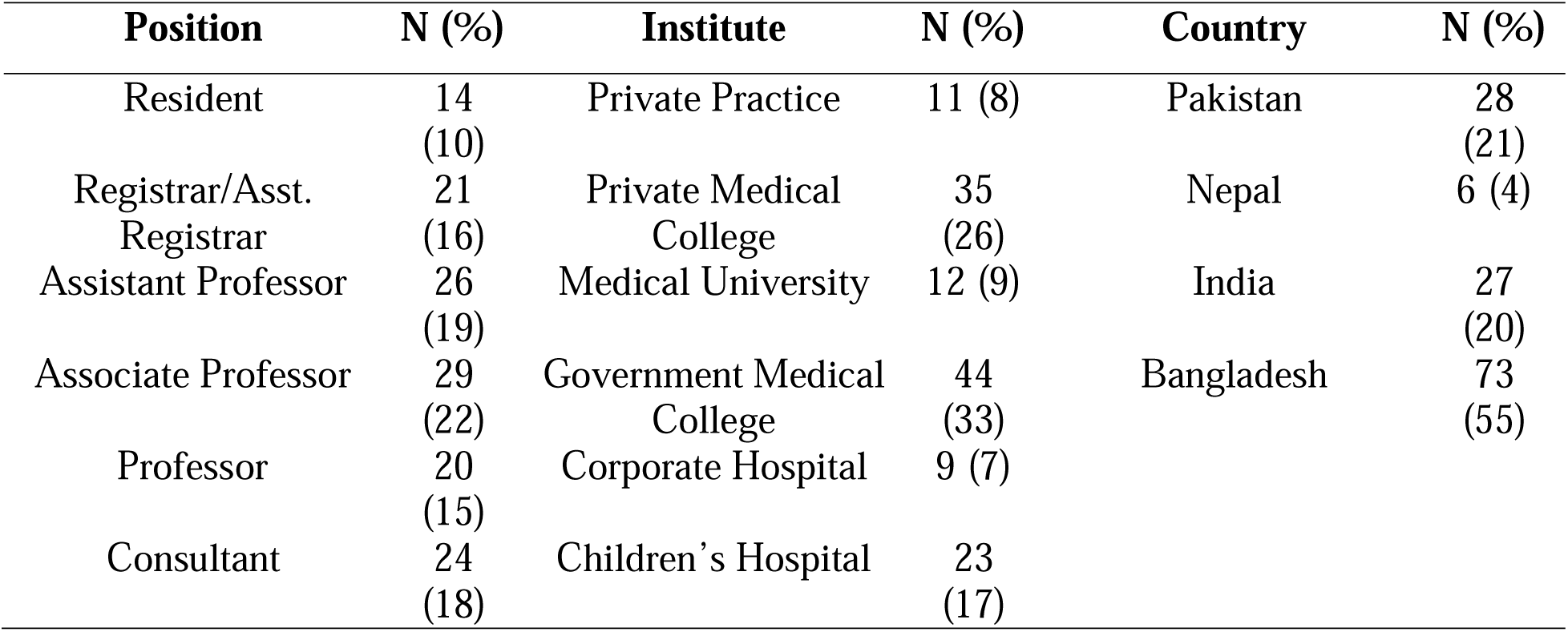
Descriptive statistic of respondents to survey summarizing distribution of position, hospital types and country of practice.

| Position | N (%) | Institute | N (%) | Country | N (%) |
| --- | --- | --- | --- | --- | --- |
| Resident | 14<br>(10) | Private Practice | 11 (8) | Pakistan | 28<br>(21) |
| Registrar/Asst.<br>Registrar | 21<br>(16) | Private Medical<br>College | 35<br>(26) | Nepal | 6 (4) |
| Assistant Professor | 26<br>(19) | Medical University | 12 (9) | India | 27<br>(20) |
| Associate Professor | 29<br>(22) | Government Medical<br>College | 44<br>(33) | Bangladesh | 73<br>(55) |
| Professor | 20<br>(15) | Corporate Hospital | 9 (7) |  |  |
| Consultant | 24<br>(18) | Children's Hospital | 23<br>(17) |  |  |

The results from the Wilcoxon signed rank tests are given in Table 2 for the entire cohort as well as grouped by position, institute, and country. Not surprisingly, there was a significant increase in the percentage of appendicitis treated non-operatively from April 1 – August 31, 2020 compared to the same time period in 2019 (P < .0001). No significant differences were found in the group analysis by institute (P = .645 within pairs; P = .516 among pairs) or country (P = .652 within pairs; P = .516 among pairs). When grouping by position, a weak significance within pairs (comparing the differences between pairs in a group) was found (P = .059) but not among pairs (P = .123), which compares the averages of the pairs between groups.

**Table 2:**
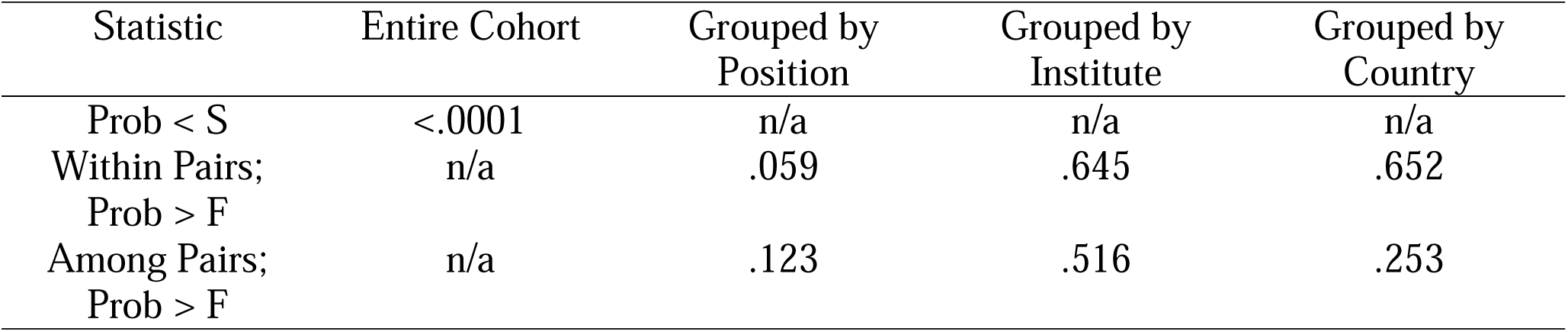
Summary of statistics from the matched pair analysis for the entire cohort and the grouped analyses by position, institute and country.

This potentially interesting effect of the position grouping is shown in more detail in Figure 1, which is the matched pair plot where the difference between pairs is plotted on the y-axis and the average between pairs is plotted on the x-axis. As this is grouped data, the y-axis reports the mean of the differences between the groups, and the y-axis represents the mean on the means. For clarity, the data points have been removed and only the group labels are plotted. The dotted red lines indicate the 95% confidence levels and the solid line the mean for the entire cohort. It should be noted that the upper 95% confidence interval falls below zero. This clearly shows that the percentage of appendicitis cases treated non-operatively has significantly increased in 2020. The grouping data suggest that the change has not been as significant for those occupying the registrar/assistant registrar position, while professors, associate professors, and consultants have treated more cases non-operatively relative to the entire cohort compared to the same time period in 2019.

**Figure 1:**
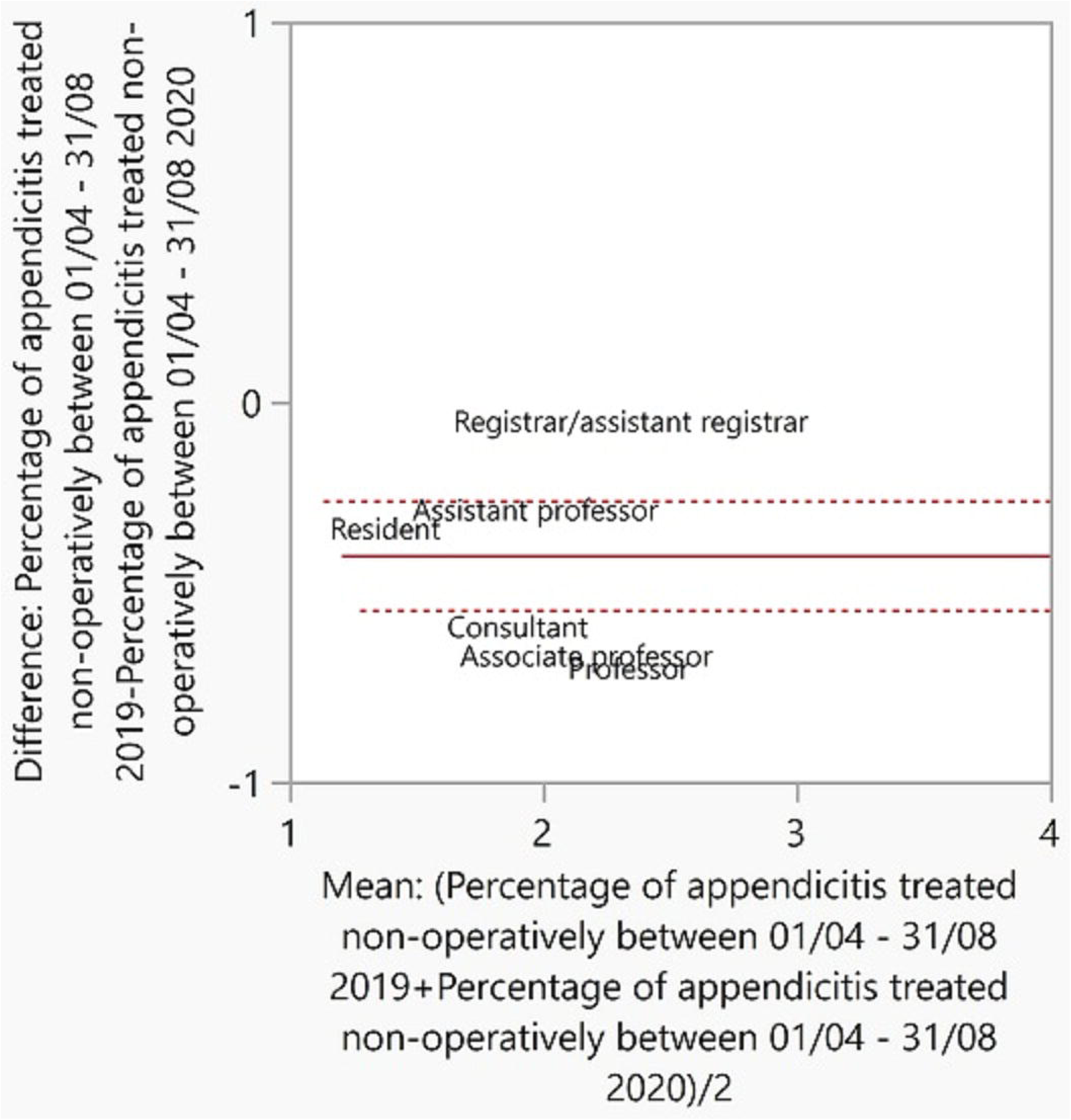
Matching pair plot showing difference between pairs is plotted on the y-axis and the average between pairs is plotted on the x-axis.

Also interesting are the results from the final question as to whether or not the physicians would consider maintaining the COVID imposed level of NOT of appendicitis after the effects of the pandemic on surgery capability subside. An interesting result from this points to the level and spirit of opinion on the topic of NOT in general. Though the respondents were given the option of replying “Don’t know” or “No comment”, the great majority (94%; 126/134) responded with a definitive “Yes” or “No”. The fractions of those grouped by position, institute, and country are shown in the mosaic plots in Figure 2. As in the matched pair analysis, no association was found using Fisher’s exact test between institute (P = .987) or country (P = .367) and a planned change in level of NOT practice after the effects of the pandemic subside. However, a significant association was found for position (P = .037). The results suggest that professors are most likely to adapt to using NOT more regularly in the post-COVID world.

**Figure 2:**
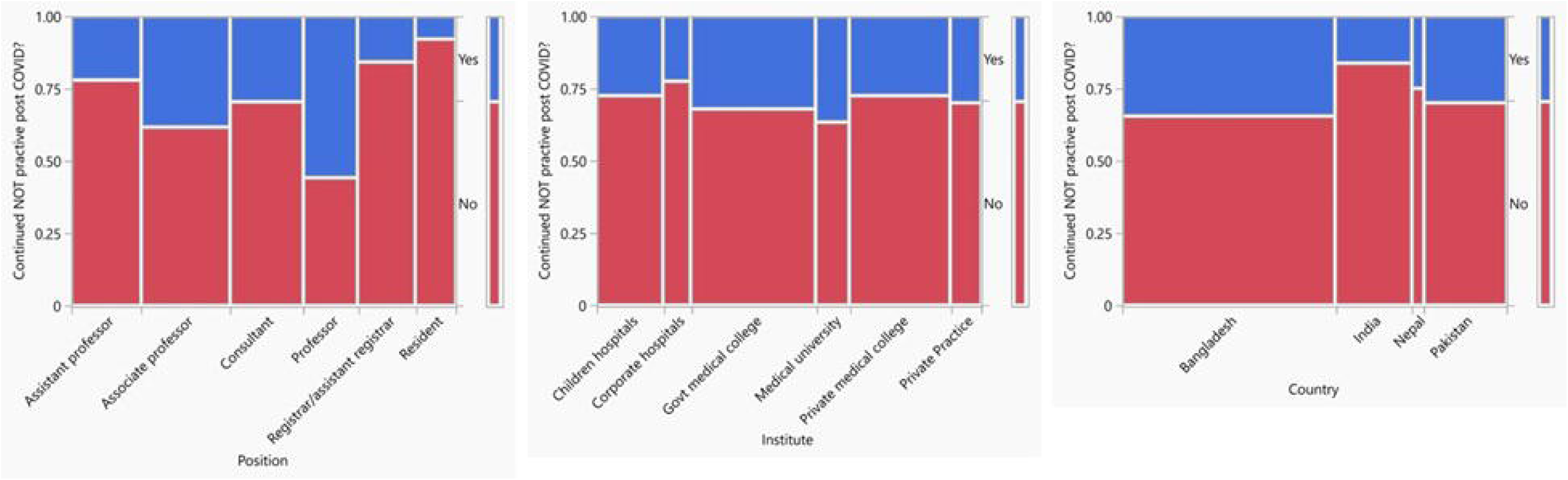
Mosaic plots illustrating distribution of responses to the question of adopting NOT of appendicitis used between April 1 and August 31, 2020,

## DISCUSSION

The results indicate that despite being required to be more reliant on usage of NOT for pediatric appendicitis necessitated by the effects of the COVID pandemic to health care, it has done little to sway the opinion of most physicians in Bangladesh, Nepal, India, and Pakistan in its place in the treatment of AUA. Only those practicing with appointed professorships have appeared to give the idea of making the increased usage of NOT a permanent practice after the effects of the pandemic wanes. The results of this study indicated an increased usage of NOT, particularly by professors, associate professors, and consultants. This is in good agreement with a recent study by Irish surgeons, where 76% of the 161 participants disclosed that they had modified their practice to a predominantly NOT approach.[21] Similarly, in a New York hospital that traditionally favors operative management, NOT usage increased to roughly 50% of all cases, with favorable results as a result of the pandemic.[22]An Indian study by Verma et al.[23] documented how their institute increased their percentage of NOT to 69% of patients, a significant increase from 22% recorded during the same time period in 2019. The recent work of Ielpo et al.[24] also showed an increased usage of NOT in pediatric appendicitis during the pandemic. However, a global study on the effect of COVID on pediatric surgery in general shows less of an effect, where only 4 out of 20 institutes were found to begin using NOT of appendicitis, and 14 out of 20 reported that they had not changed their extent of NOT.[25]

In all of these studies, little, if anything is discussed about how the physicians would modify their practice of NOT once the crisis is over. This study directly addresses this question and the results suggest that it is mainly professors who would consider adapting to change. The limitation of this study, which is independent of physicians’ personal opinions or preferences, is that the resources and capacities of hospitals will be different, a factor that was not measured in the survey. This could also sway the usage of NOT towards those facilities with the scarcest of capacities and resources. Ultimately, all of these factors can and should contribute to treatment decisions.[26]Even when the effects of the pandemic subside, the economic effects will lag considerably and recovery will be slow. This will, in turn, continue to place economic pressures on institutions that may necessitate longer than anticipated usage of NOT of AUA.

## CONCLUSIONS

The global COVID pandemic has resulted in severe disruption of surgeries worldwide, necessitating new approaches to maintain care for those in need when so many resources are being repurposed to address the massive influx of patients stricken by the virus. In the specific area of pediatric appendicitis, there is ample evidence that more hospitals and institutes are increasing their implementation of NOT of AUA in Bangladesh, India, Pakistan, and Nepal. In some respects, pediatric surgeons could think of this as their participation in an involuntary clinical trial. The results suggest that only professors in these countries would consider maintaining this increased level of practice in the post-COVID world and that the effect of hospital type or country was insignificant.

## Data Availability

The datasets generated during and/or analyzed during the current study are available at https://doi.org/10.6084/m9.figshare.13049630.v3

## ACKNOWLEDGMENTS

None

## AUTHOR CONTRIBUTIONS

All five authors designed the study, performed the experiments, analyzed the data, and wrote the manuscript.

## ETHICS STATEMENT

The study was approved by the Ethics Committee of South Point Hospital (No. Admn/SPH/190/2020).

